# Poor correlation between antibody titers and neutralizing activity in sera from SARS-CoV-2 infected subjects

**DOI:** 10.1101/2020.07.10.20150375

**Authors:** Elena Criscuolo, Roberta A. Diotti, Marta Strollo, Serena Rolla, Alessandro Ambrosi, Massimo Locatelli, Roberto Burioni, Nicasio Mancini, Massimo Clementi, Nicola Clementi

## Abstract

Plenty of serologic tests for SARS-CoV-2 have been developed so far, thus documenting the importance of evaluating the relevant features of the immune response to this viral agent. The performance of these assays is currently under investigation. Amongst them, LIAISON® SARS-CoV-2 S1/S2 IgG by DiaSorin and Elecsys Anti-SARS-CoV-2 cobas® by Roche are currently used by laboratory medicine hospital departments in Italy and many other countries. In the present study, we have firstly compared two serologic tests on serum samples collected at two different time points from forty-six laboratory-confirmed COVID-19 subjects. Secondly, eighty-five negative serum samples collected before the SARS-CoV-2 pandemic were analyzed. Thirdly, possible correlations between antibody levels and the resulting neutralizing activity against a clinical isolate of SARS-CoV-2 were evaluated. Results revealed that both tests are endowed with low sensitivity on the day of hospital admission, which increased to 97.8 and 100% for samples collected after 15 days for DiaSorin and Roche tests, respectively. The specificity of the two tests ranges from 96.5 to 100%, respectively. Importantly, a poor direct correlation between antibody titers and neutralizing activity levels was evidenced in the present study.

## Introduction

Tracking the seroprevalence of SARS-CoV-2 positive subjects certainly represents an urgent epidemiological need for facing COVID-19 pandemic waves worldwide. Additionally, commercially available serological assays are of pivotal importance from a diagnostic point of view, especially in those subjects with a clinical picture suggestive for COVID-19 but lacking a molecular-based confirmation of SARS-CoV-2 infection (1). To date, several studies have characterized the kinetics of SARS-CoV-2-elicited antibodies evidencing IgM within five days from symptoms onset for and IgG within approximatively seven days (2-5). To date, a plethora of serologic tests are invading the market, and some of them have been evaluated (6). However, other assays deserve further analyses since there is no consensus so far on antigens used for the antibody testing nor for the antibody isotype to be detected. These last aspects can be of pivotal importance for evaluating antibody response to candidate vaccines, for selecting plasmas for clinical trials, and to dissect unknown immunological aspects related to seroconversion. Indeed, as of 8^th^ July 2020, no correlations between seroconversion, neutralizing activity, and immunity have been made (7). In the present study, we evaluated the performances of two commercial serology tests, the LIAISON® SARS-CoV-2 S1/S2 IgG by DiaSorin and Elecsys Anti-SARS-CoV-2 cobas® by Roche, on 46 COVID-19 patients and 85 sera collected before the current pandemic. Data obtained from both commercial assays were then compared to the inhibitory activity of each serum against a clinical isolate of SARS-CoV-2, a clinical isolate from a patient admitted to San Raffaele Hospital, Milan (Italy) during the early COVID-19 pandemic in Lombardy.

## Materials and Methods

### Clinical Samples

The study was reviewed and approved by San Raffaele Hospital IRB in the COVID-19 Biobanking project “COVID-BioB” N° CE: 34/int/2020 19/ March/2020 ClinicalTrials.gov Identifier: NCT04318366. Forty-six serum samples were randomly collected from laboratory-confirmed symptomatic COVID-19 patients on their admission to the hospital (T0) and 15 days later (T15). Eighty-five “pre-pandemic” serum samples, spanning from 2012 to 2018, were also tested for the presence of anti-SARS-CoV-2 antibodies.

### Immunoassays

Elecsys Anti-SARS-CoV-2 cobas® by Roche and LIAISON® SARS-CoV-2 S1/S2 IgG assay by DiaSorin were used for detecting anti-SARS-CoV-2 antibodies in all serum samples. Analyses were performed according to manufacturer’s instruction by using cobas® and LIAISON® XL Analyzer platforms. In brief, Elecsys by Roche uses a recombinant SARS-CoV-2 nucleocapsid (N) antigen. The electrochemiluminescence immunoassay (ECLIA) can detect the presence of IgG, IgM, and IgA antibodies recognizing the N protein (8). According to the producer, samples positive for anti-SARS-CoV-2 antibodies show a cutoff index (COI) equal to or greater than 1. All samples with a COI < 1.0 are considered negative for the presence of SARS-CoV-2 antibodies. The SARS-CoV-2 S1/S2 IgG assay by DiaSorin can detect IgG antibodies directed against two recombinant SARS-CoV-2 proteins: the S1 and S2 which are involved in both docking and fusion processes of the virus (9). According to manufacturer instructions, the test by DiaSorin can detect the presence of neutralizing antibodies directed against the spike protein of SARS-CoV-2. Samples featuring < 12.0 AU/mL are considered negative according to manufacturer instructions, those ranging between 12.0 to 15.0 AU/mL are undetermined and those above 15 AU/mL are positive.

### Virus and Cells

Vero E6 (Vero C1008, clone E6 – CRL-1586; ATCC) cells were cultured in Dulbecco’s Modified Eagle Medium (DMEM) supplemented with non-essential amino acids (NEAA), penicillin/streptomycin (P/S), Hepes buffer and 10% (v/v) Fetal bovine serum (FBS). A clinical isolate of SARS-CoV-2 (hCoV-19/Italy/UniSR1/2020; GISAID Accession ID: EPI_ISL_413489) was obtained and propagated in Vero E6 cells.

### Virus titration

Virus stocks were titrated using both Plaque Reduction Assay (PRA, PFU/mL) and Endpoint Dilutions Assay (EDA, TCID_50_/mL). In PRA, confluent monolayers of Vero E6 cells were infected with eight 10-fold dilutions of virus stock. After 1 h of adsorption at 37°C, the cell-free virus was removed. Cells were then incubated for 48 h in DMEM containing 2% FBS and 0.5% agarose. Cells were fixed and stained, and viral plaques were counted. In EDA, Vero E6 cells were seeded into 96 wells plates and infected at 95% of confluency with base 10 dilutions of virus stock. After 1 h of adsorption at 37°C, the cell-free virus was removed, cells were washed with PBS 1X and complete medium was added to cells. After 48h, cells were observed to evaluate the presence of cytopathic effect (CPE). TCID_50_/mL of viral stocks were then determined by applying the Reed-Muench formula.

### Microneutralization experiments

Vero E6 cells were seeded into 96 wells plates 24 h prior to the experiment performed at 95% cell confluency for each well. Decomplemented serum dilutions (1:100, 1:200, 1:400, and 1:800) were incubated with SARS-CoV-2 at a 0.001 multiplicity of infection (MOI) for 1 h at 37°C. Virus-serum mixtures and positive infection control were applied to Vero E6 monolayers after washing cells with PBS 1X, and virus adsorption was carried out at 37°C for one hour. Then, cells were washed with PBS1X to remove cell-free virus particles and virus-containing mixtures and controls were replaced with complete DMEM supplemented with 2% FBS. Plates were incubated at 37°C on the presence of CO_2_ for 72 h. The experiments were performed in triplicate. Neutralization activity was evaluated by comparing CPE presence detected in the presence of virus-serum mixtures to positive infection control.

### Data analysis and statistics

Test performances were evaluated by sensitivity and specificity with the associated standard error (SE) and 95% confidence interval (CI). Differences between sensitivities and specificities, respectively, were assessed by exact binomial test for paired study design (10). The overall performances were also measured, by means of the ROC curves and the associated Area Under the Curve (AUC). For graphical purposes, we considered smooth ROC curves based on kernel estimators (11) with unbiased cross-validation bandwidth selection. We verified differences between AUCs by means of stratified bootstrap resampling for paired data (12). Relationships between values obtained with the two diagnostic methods and neutralizing activity of sera were investigated by Spearman’s correlation coefficient and fitting spline functions. P-values lower than *P* = 0.05 were considered significant. Al computation and analysis were performed in the R environment (R ver. 4.0.0).

## Results

### Sensitivity and specificity of DiaSorin and Roche diagnostic tests

Pre-pandemic sera were tested with DiaSorin and Roche diagnostic assays (**Fig. 1A**). Three out of 85 samples tested positive with DiaSorin, and one tested in the “grey-zone” (12.0-15.0 AU/mL) with the same test. Thus, the diagnostic specificity observed on the tested samples for the DiaSorin assay was 96.5% (SE: 2%; 95% CI: 92.5–100%) (**Fig. 1C**). Then, forty-six serum samples were randomly collected from laboratory-confirmed symptomatic COVID-19 patients. Admission to the hospital (T0) and 15 days later (T15) time points were evaluated for each patient, showing an overall increase in serum IgG titers 15 days after hospital admission (**Fig. 1B)**. The diagnostic sensitivity at T0 was 19.6% (SE: 5.8%; 95% CI: 8.1–31%), and at T15 was 100% (SE: 0%; 95% CI: 100-100%). On the other hand, the diagnostic specificity of the Roche test was 100% (SE: 0%; 95% CI: 100–100%) and its diagnostic sensitivity at T0 was 45.7% (SE: 7.3%; 95% CI: 31.2–60%), and at T15 was 100% (SE: 0%; 95% CI: 100-100%). Roc curves, calculated for the two tests at the two different time points, show comparable performance at T15 (*P* = 0.2961). Thus, Roche diagnostic test showed a statistically better performance than DiaSorin at T0 on the tested samples (*P* < 0.001) (**Fig. 1D**).

**Fig. 1.**
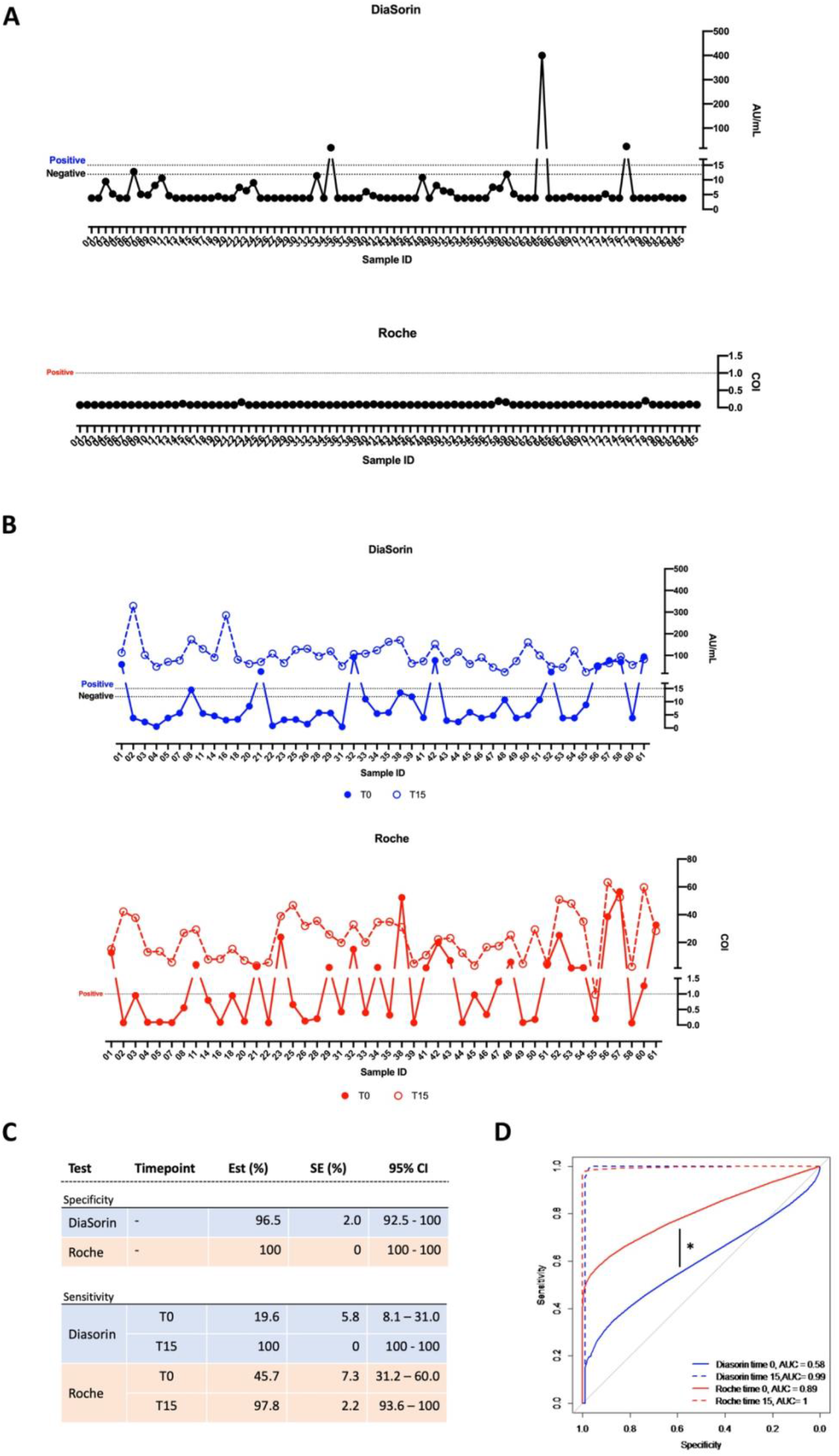
Sensitivity and specificity of the two commercial assays. **A)** “Pre-pandemic” samples tested with both DiaSorin and Roche assays (●, black line). **B)** Antibody levels detected by DiaSorin (●, T0 solid blue line, T15 dotted blue line) and Roche (●, T0 solid red line, T15 dotted red line) tests of sera from subjects with positive nasopharyngeal swabs. **C)** Sensitivity and specificity observed on the tested samples at T0 and T15 are reported for the two diagnostic tests. Standard error (SE) and 95% confidence interval (CI) for all values are also reported. **D)** Roc curves for DiaSorin (T0 solid blue line, T15 dotted blue line) and Roche (T0 solid red line, T15 dotted red line) tests are reported. * *P* < 0.001

### Neutralizing activity evaluation of a limited cohort of serum samples

Five patients were randomly selected for the characterization of the neutralizing activity of their serum samples against SARS-CoV-2, at both T0 and T15 (**Fig. 2**). Results showed that samples collected at T0 were not able to neutralize virus infection, consistent with the low antibody titers detected by both diagnostic tests. At T15, only one out of five samples (ID #4) showed detectable neutralizing activity also when a high dilution (1:800) was used, while two samples (ID #2 and #3) strongly neutralized the infection only when used at 1:100 with a very low neutralizing activity, 22.2% for sample ID #2 and 11.1% for sample ID #3, still detectable when used 1:200. Notably, the T15 of sample ID #4 potently inhibited viral replication despite its low antibody titer compared to other T15-samples detected by DiaSorin and Roche (48.1 AU/mL and 13 COI respectively). Finally, serum from patient #2 showed complete neutralizing activity at low dilution (1:100) and 22.2% neutralizing activity at 1:200.

**Fig. 2.**
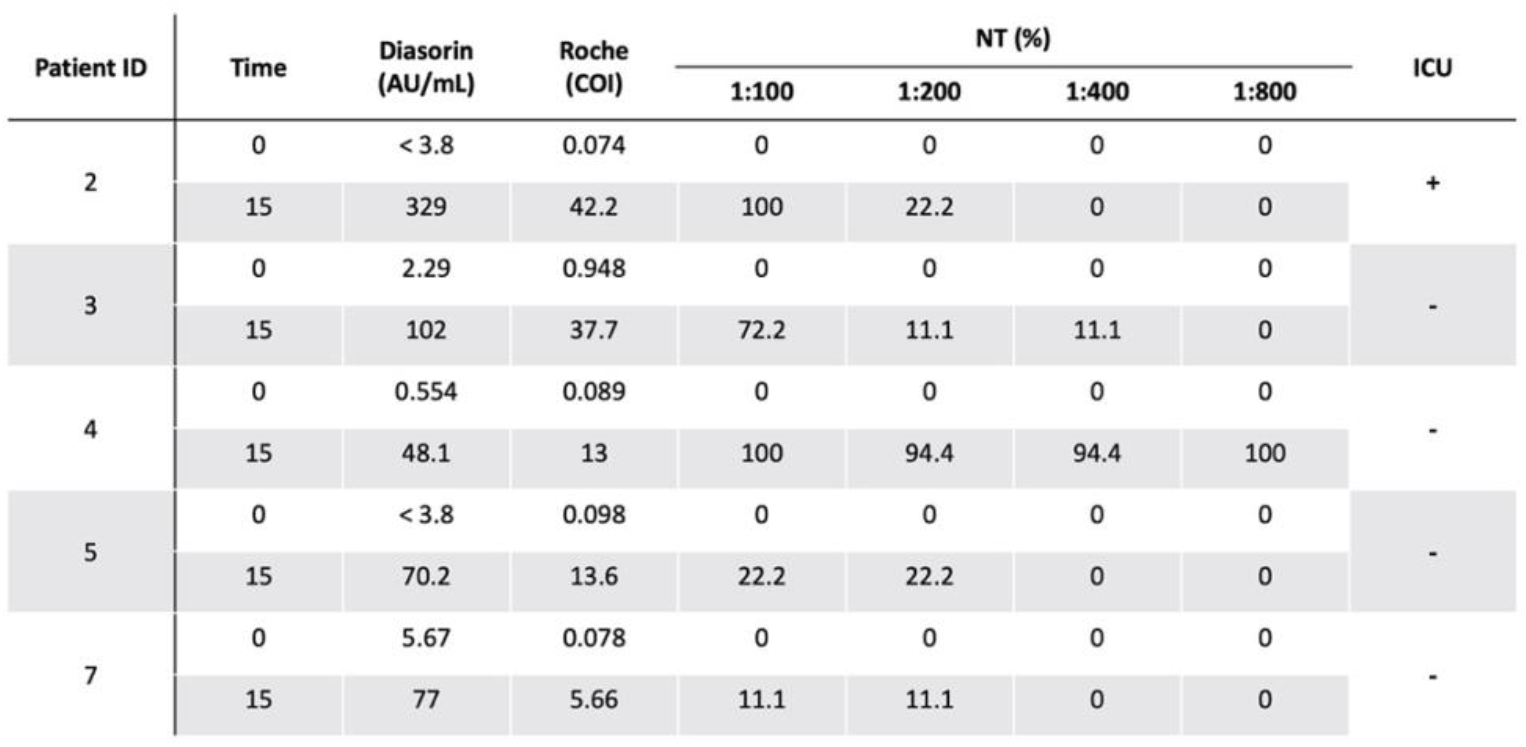
Evaluation of serum neutralizing activity at four different serum concentrations. Neutralizing activity of five patients’ serum, at T0 and T15. Sera were diluted 1:100, 1:200, 1:400, and 1:800 and tested with 0.001 MOI of SARS-CoV-2. Antibody titers detected for each serum at both timepoints are also reported for DiaSorin and Roche, respectively. One out of five patients admitted to ICU is highlighted in the graph.

### Neutralizing activity evaluation of all tested sera

As the previous analysis showed that no neutralizing activity was detected at T0, all remaining sera were tested at T15. Moreover, all T15 samples were tested at a 1:200 dilution, based on what observed in the five sera tested at different dilutions. Sera neutralizing activity does not directly correlate with antibody titers detected with both DiaSorin and Roche test, as highlighted by the graph (**Fig. 3A**). Moreover, no apparent relationship between ICU admission and both antibody titer and neutralizing activity was observed. None of the five “pre-pandemic” sera checked for their neutralizing activity, including those testing positive with the DiaSorin kit, neutralized the virus. Spearman correlation analysis confirms the lack of correlation between anti-SARS-CoV-2 antibody presence detected with both diagnostic serologic assays and the neutralizing activity (**Fig. 3B**).

**Fig. 3.**
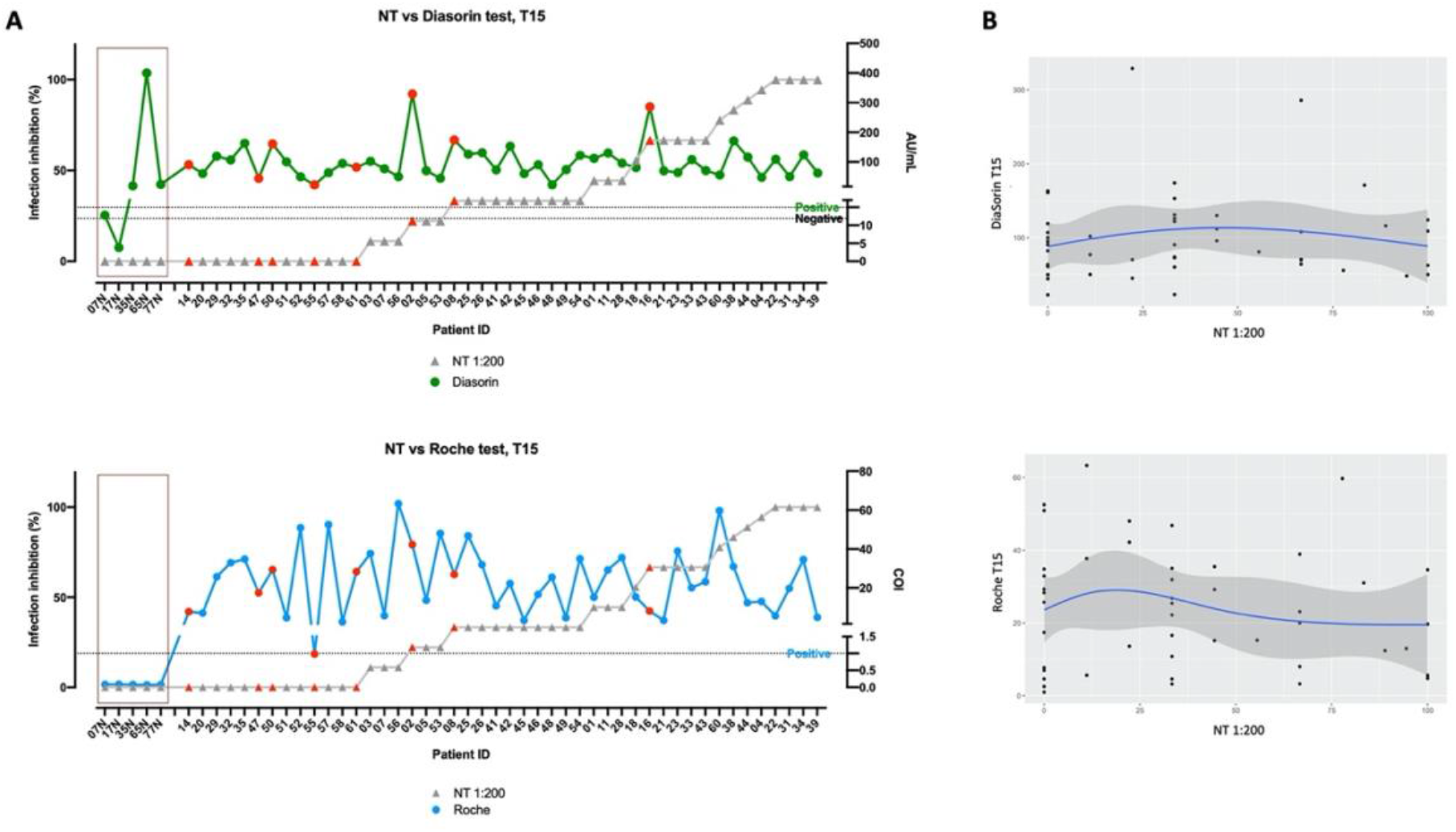
Characterization of anti-SARS-CoV-2 serum antibodies and evaluation of neutralizing activity. **A)** graphs rank the sera basing on their neutralizing capability. All sera were tested at a dilution of 1:200 (▴, grey line). Levels of SARS-CoV-2 specific antibodies detected with the tests by Roche and DiaSorin are also indicated (●, green and blue lines). Red dots indicate patients admitted to ICU. **B)** Spearman correlation analyses between values obtained with the two diagnostic methods and neutralizing activity of sera.

## Discussion

A comparative analysis between two serologic assays for the detection of antibodies directed against SARS-CoV-2 was carried out on sera collected from subjects testing positive for SARS-CoV-2 RNA on nasopharyngeal swabs. The same analyses were also performed on sera collected before the COVID-19 current pandemic. Overall, both commercial assays are characterized by a good sensitivity when analyzing serum samples collected from subjects 15 days after their presentation to the physician (T15), whit test by DiaSorin behaving slightly better compared to Roche at this timepoint. Even so, LIAISON SARS-CoV-2 S1/S2 by DiaSorin showed less specificity than the Roche test since it detected anti-SARS-CoV-2 antibodies also on control sera collected well before the COVID-19 pandemic. What observed for positive results obtained with DiaSorin assays on “pre-pandemic” sera was possibly due to cross-reactions in accord to what described by the producer which reported 3 out of 168 positive detections in sera collected before October 2019 (9). The sensitivity of both DiaSorin and Roche assays was dramatically lower (19.6% and 45.7% respectively *P*<0.001), for sera collected at T0. Differences in terms of sensitivity between the two methods at T0 could be mainly attributed to the capability of Roche assay to detect also for IgM antibodies (8). This observation can be of help when testing serology in subjects with a clinical picture suggestive for COVID-19 but negative to the direct detection of SARS-CoV-2 on nasopharyngeal swabs or bronchoalveolar lavages. Other important differences for the two commercial assays possibly impacting their performances include the SARS-CoV-2 recombinant antigens included in the commercial kits. The Elecsys by Roche detects antibodies able to selectively bind a recombinant form of nucleocapsid protein N of SARS-CoV-2. Whereas, DiaSorin assay detects IgGs able to bind the S1 or the S2 recombinant portions of the virus spike glycoprotein. The S protein mediates at least two crucial steps in the early phases of the viral productive infection: the docking to host cell receptor or putative co-receptors and the fusion process. This observation and the need to infer possible correlations between the presence of antibodies against SARS-CoV-2 S protein triggered the second round of laboratory investigations we performed. For this purpose, all samples from COVID-19 subjects already tested with both serological commercial tests were also tested for their neutralizing activity against a clinical isolate of SARS-CoV-2. A first pilot microneutralization experiment was performed by using 0.001 MOI of the virus against several dilutions of a small cohort of five sera. From this experiment, it was evident that the detected antibody levels were unrelated to the neutralizing capability of sera tested at dilutions spanning from 1:100 to 1:800. On that basis, all sera were then checked for their neutralizing activity against a clinical isolate of SARS-CoV-2 at a dilution of 1:200. Results underline the lack of relationship between antibody titer detected with the two commercial tests and neutralizing activity. This observation does not surprise for antibodies directed against N proteins revealed by Roche test but should cautiously be considered when referred to the indication reported by the DiaSorin brochure of the commercial kit which states that the test can give a clue on the presence of neutralizing antibodies directed against SARS-CoV-2 (9). Even more importantly this observation can impact directly on the methods to be used for detecting possible correlates of protection from infections for subject enrolled in vaccine clinical trials based on S protein (13, 14). However, it is of pivotal importance to highlight that even the serum neutralizing activity was not related to protection so far. Gathering our observations and literature data we believe that great efforts are still necessary for implementing observations on antibody kinetics in order to develop novel diagnostic algorithms useful both for epidemiological and clinical purposes. Moreover, further investigations elucidating the clinical role of neutralizing antibodies and the possibility of detecting them with binding assays will be of paramount importance for addressing the development of effective vaccines.

## Data Availability

The datasets generated during and/or analyzed during the current study are available from the corresponding author on reasonable request.

